# Outcome of community onset severe sepsis, Sepsis-3 sepsis, and bacteremia in Sweden – a prospective population-based study

**DOI:** 10.1101/2024.04.18.24306009

**Authors:** Lars Ljungström, Rune Andersson, Gunnar Jacobsson

## Abstract

**Background:** Register-based studies are common in sepsis epidemiology. Chart review is considered gold standard but is time consuming. This is one of few such studies.

**Methods:** In a 9-month prospective and consecutive study conducted in 2011-12, chart review was used to investigate outcomes in patients with severe sepsis, Sepsis-3 sepsis, and bacteremia in a population of 256,700 inhabitants in southwest Sweden. All adult patients aged ≥18 years admitted to hospital and given intravenous antibiotic treatment within 48 hours were evaluated, N=2,196. Cohort mortality was calculated up to 10 years after admission.

**Results:** Among 2,072 adults with any infection, 429 patients had severe sepsis of which 59 had septic shock. The 28-day case fatality rate (CFR) was 25%, 41% in those with septic shock. Sepsis-3 sepsis was diagnosed in 1,299 patients. The 28-day CFR was 12%. Among the 1,299, 393 also had severe sepsis. In 906 patients with Sepsis-3 sepsis but not severe sepsis, the 28-day CFR was 6%. For both sepsis definitions, the 28-day CFR increased 10-fold between the youngest and the oldest age groups. Age >75 years, and renal dysfunction were the strongest independent risk factors for 28-day case fatality. Bacteremia occurred in 283/2,072 (13%) patients. The 28-day CFR was 13% overall, 25% in severe sepsis and 4% in non-severe sepsis. Even 10 years after admission, the mortality rate was higher in sepsis patients by either definition.

**Conclusions:** The 28-day case fatality rate (CFR) in patients with Sepsis-3 sepsis, 12%, is the result of a large group of patients with a low 28-day CFR, 6%, camouflaging a group with severe sepsis and a high 28-day CFR, 25%. Age >75 years is an independent risk factor for case fatality. The 28-day CFR in patients with bacteremia is a function of severe sepsis, not bacteremia *per se*. Even after ten years, mortality is increased in both sepsis groups.

## Introduction

In an American consensus meeting in 1991, sepsis (Sepsis-1) was defined as the presence of two of four criteria for the systemic inflammatory reaction syndrome (SIRS), if caused by an infection (1). Sepsis with organ dysfunction was called “severe sepsis”, but organ dysfunction was not clearly defined. A second international consensus meeting on sepsis definitions in 2001 (Sepsis-2), expanded the criteria for sepsis, but could not present unanimous criteria for severe sepsis either (2). Thus, similar yet different criteria for organ dysfunction in severe sepsis have been used in studies up until 2016, and in some cases later. In 2011, a work group within the Swedish Infectious Disease Society published criteria for severe sepsis and septic shock that were used in this study (3) (S 1 Table). These are similar to the criteria presented in 2012 by the Surviving Sepsis Campaign (4). The main difference from mostly used criteria for organ dysfunction in sepsis studies is the stricter criteria for respiratory dysfunction. In the Swedish definition, fulfilling two out of four SIRS criteria could be replaced by “documented infection”. The Sepsis-2 definition of septic shock in adults was severe sepsis together with persistent arterial hypotension. Hypotension was defined as “systolic arterial pressure below 90 mmHg, mean arterial pressure lower than 60, or a reduction in systolic blood pressure of more than 40 mmHg from baseline, despite adequate volume resuscitation” (2).

In 2016 a new sepsis definition, commonly called Sepsis-3, was launched. Sepsis is now defined as “organ dysfunction caused by a dysregulated host response to infection” (5). The criterion for sepsis according to Sepsis-3 is a 2-point increase from a patient’s base line values in the Sequential Organ Failure Assessment (SOFA) score. At the same time, septic shock was redefined. The clinical criterion in adult patients is now “hypotension requiring vasopressor therapy to maintain a mean arterial blood pressure (MAP) of 65 mm Hg or greater and having a serum lactate level greater than 2 mmol/L after adequate fluid resuscitation” (6).

For studies on sepsis epidemiology, chart review is considered gold standard (7). Chart reviews are time consuming and are rarely used for whole populations. In this 9-month prospective study in 2011-2012, chart review was used to investigate the incidence of community onset severe sepsis in a well-defined population of 256,700 inhabitants in southwest Sweden. The incidence was 279/100,000/year. After the new Sepsis-3 definition was published in 2016, the Sepsis-3 criteria were retrospectively applied to this cohort. The incidence of community onset Sepsis-3 sepsis was 846/100,000/year. The differing incidences were mainly due to the milder criteria for respiratory dysfunction used in Sepsis-3 (8). In Sepsis-3 the criterion for lung dysfunction corresponds to a saturation level of ≤91% in otherwise healthy patients, but in severe sepsis it is ≤86% or ≤78% if the lung is the focus of the infection.

Both severe sepsis and Sepsis-3 sepsis are associated with high case-fatality rates (CFR) ranging between 5-55% (9–11). The large variation can be explained by differences in methods of data abstraction, populations studied, criteria used, definitions of infection, and application of these variables (12).

Bacteremia, often referred to as “blood stream infection”, is highly associated with sepsis, having a CFR of approximately 10-15% (13, 14). Bacteremia studies are mostly retrospective, performed on laboratory databases of positive blood cultures, and can therefore be said to represent bacteremia in whole populations. On the other hand, data on organ dysfunction is generally not available, so the relationship between CFR and organ dysfunction has not been evaluated comprehensively.

Among factors affecting outcome, early appropriate antibiotic treatment is said to be of key importance (15). This has been shown for patients with septic shock (16), but is more widely debated concerning patients with severe sepsis.

Increasing age is a risk factor for case fatality in sepsis patients (17). Some studies even show age to be an independent risk factor for sepsis-related CFR (18–20), but the age factor is not often discussed in sepsis studies.

Elevated body temperature is one of the hallmarks of infection. Yet not all patients with sepsis are febrile at initial presentation. Low temperature on arrival has been described as an independent risk factor for case fatality in patients with bacteremia (21) and in patients with severe sepsis treated in the ICU (22, 23).

Long-term mortality is increased following sepsis, indicating that sepsis may cause lasting injury to the individual even several years after the insult (24–26).

The aim of this study was to describe short-term outcomes in patients with severe sepsis, Sepsis-3 sepsis, and bacteremia in this population. We also investigated risk factors for case fatality, focusing on time to antibiotic treatment, age, and temperature at presentation. Finally, we briefly describe the long-term mortality in those who had sepsis by either definition compared to those who did not have sepsis.

## Methods

The original study was approved by the Ethical Review Board at the University of Gothenburg (permit 376/2011). The 10-year follow up was approved by the Swedish Ethical Review Authority (2022-04814-02).

In a 9-month prospective and consecutive study chart review was used to investigate the incidences of community onset severe sepsis, Sepsis-3 sepsis, and bacteremia in a 256,700 population in southwest Sweden. All adult patients aged ≥18 years admitted to the hospital between September 8, 2011 - June 7, 2012, and treated with intravenous antibiotics within 48 hours were evaluated, N=2,196 (8).

This cohort was analysed for patient risk factors and outcomes in those with severe sepsis, Sepsis-3 sepsis, and bacteremia. Data from the hospital electronic health record were used for patient risk factors. The 28-day, 1-year, and 10-year case fatality rates (CFR) were calculated by use of the unique Swedish personal identification number and the Swedish death registry. Data for septic shock could only be evaluated for septic shock using Sepsis-2 criteria. Septic shock according to sepsis-3 criteria could not be evaluated since lactate measurements at the time of the study were performed on a local instrument in the ICU and the results were not entered into the electronic health record. For bacteremia, only the 28-day CFR was calculated. Base-line data refers to the most deviating laboratory values or vital signs registered by the Emergency Medical Services (EMS) or in the Emergency Department (ED), or in the ward within 48 hours after admission, when sepsis was first suspected. Dependent as well as independent risk factors for case fatality were calculated.

### Definitions

Appropriate empirical antibiotic treatment refers to antibiotic treatment according to current national guidelines. Appropriate etiological treatment refers to antibiotic treatment according to antibiotic susceptibility pattern.

### Statistical analyses

Descriptive statistics as mean and standard deviation for continuous data and frequencies and percentages for categorical data are presented. For ordinal data or skewed distributed data median with quartiles are presented. Both parametrical and non-parametrical tests were used for univariate comparisons, depending on data type.

In a univariate analysis, age groups, empiric antibiotic treatment, time to appropriate antibiotic treatment, co-morbidities, gender, vital signs at baseline, biochemistry at baseline and individual organ dysfunctions were evaluated for association with 28-day case fatality. All variables with a p-value <0.2 in univariate comparisons were included in a multivariate regression analysis.

Logistic regression was used as model to explore in which degree severe sepsis, Sepsis-3 and other factors are associated with 28-day CFR. One- and 10-years survival was analysed, and different groups were compared by Kaplan-Meier analysis and log-rank test. Two-sided testing was used for all analyses, and a p-value <0.05 was regarded as statistically significant. Data were analysed using IBM SPSS version 25.0 (Inc, Chicago, IL).

## Results

### Baseline characteristics

Baseline characteristics in 28-day survivors versus non-survivors by either sepsis definition are displayed in **Table 1**. Organ dysfunction in Sepsis-3 sepsis was defined for each organ system as a ≥2 point increase in the SOFA score from baseline. The 173 patients with Sepsis-3 sepsis having only 1-point increases in any organ system were excluded in this calculation. In summary, non-survivors were significantly older than survivors, had significantly more organ dysfunctions and significantly more comorbidities. For baseline characteristics of the whole cohort, see S2 Table.

**Table 1.** Baseline characteristics in 28-day survivors versus non survivors having severe sepsis or Sepsis-3-sepsis respectively.

|  | Severe sepsis, N=429 |  |  | Sepsis-3 sepsis, N=1,299 |  |  |
| --- | --- | --- | --- | --- | --- | --- |
|  | Survivors<br>n=322 | Non-survivors<br>n=107 | p-value | Survivors<br>n=1,144 | Non-survivors<br>n=155 | p-value |
| Age years, mean (SD) | 73 (16) | 80 (11) | <0.001 | 73 (16) | 81 (10) | <0.001 |
| Age years, median (IQR) | 75 (65-84) | 82 (73-87) | <0.001 | 77 (66-84) | 83 (76-88) | <0.001 |
| Male sex, n (%) | 174 (54) | 57 (53) | 0.890 | 627 (55) | 81 (52) | 0.550 |
| Age-group, n (%) |  |  | <0.001 |  |  | <0.001 |
| 18-49 years | 28 (9) | 1 (1) |  | 94 (8) | 2 (1) |  |
| 50-64 years | 46 (15) | 7 (6) |  | 168 (15) | 8 (5) |  |
| 65-74 years | 79 (25) | 20 (19) |  | 249 (22) | 24 (16) |  |
| **75-84 years** | 92 (29) | 35 (33) |  | 350 (31) | 49 (32) |  |
| **≥85 years** | 77 (24) | 44 (41) |  | 283 (25) | 72 (46) |  |
| **Any co-morbidity, n (%)** | 288 (89) | 104 (97) |  | 1,031 (90) | 149 (96) |  |
| **Cardiovascular** | 200 (62) | 74 (69) | 0.189 | 713 (62) | 111 (72) | 0.024 |
| **Chronic pulmonary disease** | 56 (17) | 19 (18) | 0.931 | 236 (21) | 27 (17) | 0.351 |
| **Chronic renal disease** | 21 (7) | 8 (8) | 0.733 | 75 (7) | 12 (8) | 0.579 |
| **Chronic hepatic disease** | 1 (0.3) | 1 (0.9) | 0.412 | 6 (0.5) | 2 (1) | 0.253 |
| **Diabetes mellitus** | 78 (24) | 27 (25) | 0.833 | 227 (20) | 30 (19) | 0.886 |
| **Malignancy, any** | 50 (16) | 30 (28) | 0.004 | 198 (17%) | 43 (28%) | 0.002 |
| **Immunosuppression** | 14 (4) | 6 (6) | 0.592 | 47 (4) | 8 (5) | 0.541 |
| **No comorbidity** | 34 (11) | 3 (3) | 0.013 | 113 (10) | 6 (4) | 0.015 |
| **Average co-morbidity, mean (SD)** | 1.7 (1.0) | 2.0 (1.0) | 0.005 | 1.6 (1.0) | 1.9 (1.0) | 0.001 |
| **Bacteremia, n (%)** | 94 (29) | 29 (27) | 0.679 | 180 (16) | 31 (20) | 0.177 |
| **Organ dysfunction** |  |  |  |  |  |  |
| **Hypotension, n (%)** | 130 (40) | 49 (47) | 0.235 | 131 (20) | 48 (41) | <0.001 |
| **Hypoperfusion, n (%)** | 165 (51) | 58 (54) | 0.222 | 138 (21) | 56 (48) | <0.001 |
| **Lactate mg/L, mean (SD)** | 4.6 (1.8) | 5.5 (2.9) | 0.047 | 4.6 (1.9) | 5.5 (2.9) | 0.062 |
| **Pulmonary dysfunction, n (%)** | 103 (32) | 57 (55) | <0.001 | 104 (16) | 58 (50) | <0.001 |
| **Renal dysfunction, n (%)** | 54 (18) | 40 (44) | <0.001 | 55 (9) | 40 (39) | <0.001 |
| **Hepatic dysfunction, n (%)** | 3 (1) | 5 (5) | 0.010 | 3 (0.5) | 5 (4) | <0.001 |
| **Coagulation dysfunction, n (%)** | 30 (10) | 17 (17) | 0.037 | 30 (5) | 17 (15) | <0.001 |
| **Cerebral dysfunction, n (%)** | 82 (26) | 45 (42) | 0.001 | 157 (14) | 55 (36) | <0.001 |
| **Septic shock, n (%)** | 35 (12%) | 24 (25%) | 0.001 | n.d. | n.d. | n.d. |
| **Average dysfunctioning organs, mean (SD)** | 1.8 (1.0) | 2.5 (1.6) | <0.001 | 1.2 (0.6) | 1.5 (0.8) | <0.001 |
| **Treatment in the ICU, n (%)** | 83 (26) | 24 (22) | 0.488 | 102 (9) | 28 (18) | <0.001 |
| **Biochemistry on arrival, median (IQR)** |  |  |  |  |  |  |
| **Hemoglobin, g/L** | 130 (118-145) | 123 (111-141) | 0.016 | 130 (117-142) | 124 (108-140) | <0.001 |
| **WBC x 10<sup>9</sup>/L** | 13.1 (10-17) | 13.1 (8-18) | 0.566 | 12.3 (9-16) | 13.0 (9-18) | 0.170 |
| **Neutrophils, x 10<sup>9</sup>/L** | 11.4 (7.1-15.8) | 11.6 (5.7-17.3) | 0.830 | 10.0 (7-14) | 11.0 (6-17) | 0.106 |
| **Lymphocytes, x 10<sup>9</sup>/L** | 0.7 (0.4-1.1) | 0.8 (0.4-1.3) | 0.198 | 0.8 (0.5-1.2) | 0.8 (0.5-1.3) | 0.672 |
| **NLCR** | 16.2 (9-28) | 14.6 (6-21) | 0.039 | 11.4 (7-21) | 14.4 (7-22) | 0.465 |
| **CRP, mg/L** | 164 (102-253) | 201 (108-302) | 0.047 | 138 (80-200) | 192 (90-280) | <0.001 |
| **Lactate, mmol/L** | 3.3 (2-4) | 3.3 (2-4.6) | 0.093 | 1.9 (1.4-2.6) | 2.6 (1.9-3.8) | <0.001 |
| **Temperature, °C, mean (SD)** | 38.0 (1.2) | 37.4 (1.0) | <0.01 | 38.1 (1.1) | 37.4 (1.2) | <0.01 |
| **SIRS ≥2 n (%)** | 289 (90) | 91 (85) | 0.185 | 955 (83) | 128 (83) | 0.778 |

ICU = Intensive Care Unit; WBC = White Blood cell Count; NLCR = Neutrophil to Lymphocyte Count Ratio; SIRS = Systemic Inflammatory Response Syndrome.

### Case fatality rates in severe sepsis and Sepsis-3 sepsis

The 28-day CFR in the 2,196 individual patients in the study was 8.6% (188/2,196). In 124/2,196 of the patients (6%) no infection could be diagnosed. The 28-day CFR was 12% (15/124). In 429 patients having severe sepsis the 28-day CFR was 25% (107/429). In those with infection but not severe sepsis, it was 4% (66/1,643). **Fig 1**.

**Fig 1.**
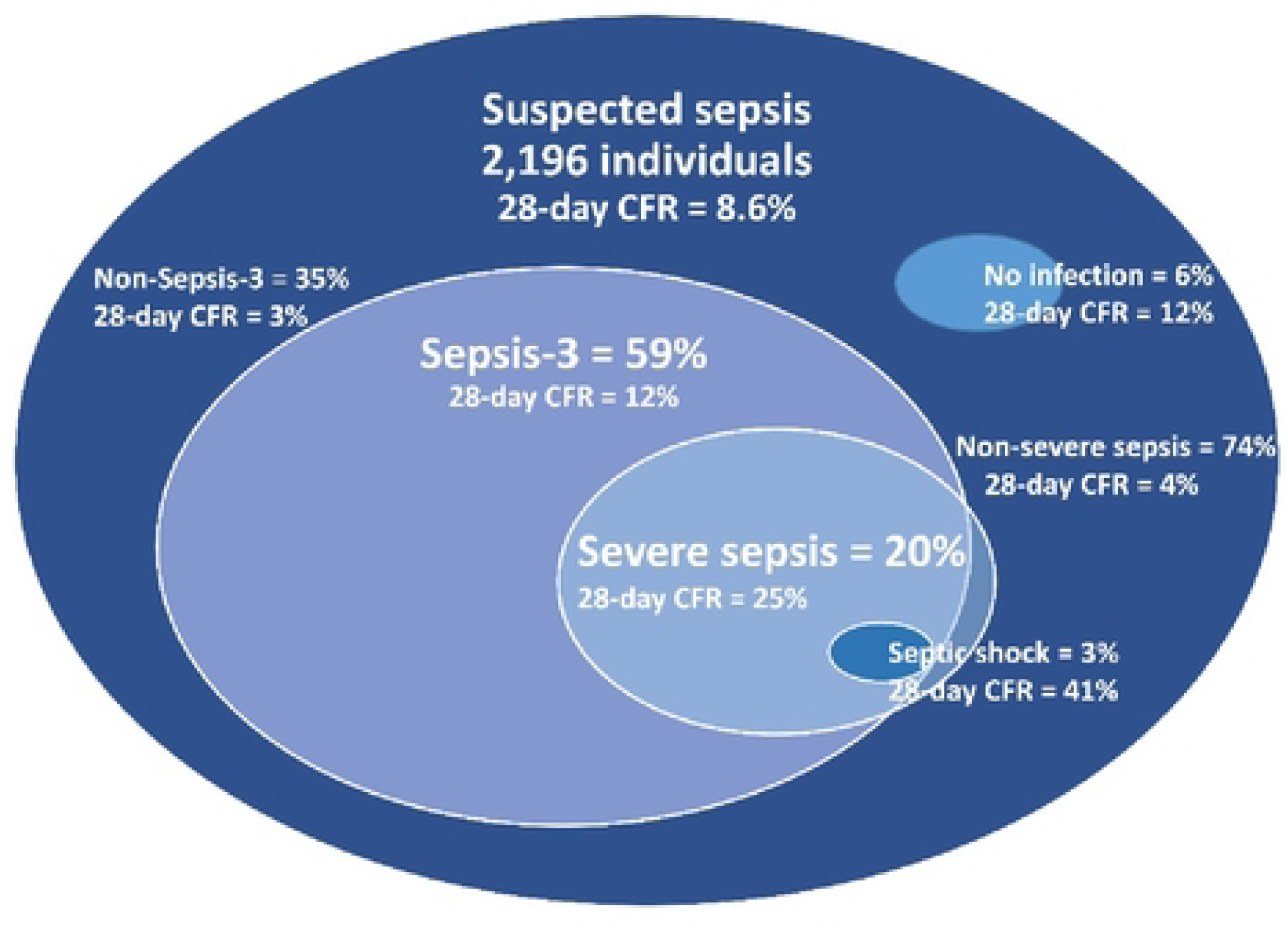
The 28-day case fatality rates in severe sepsis and Sepsis-3 sepsis within a cohort of 2,196 consecutive patients with suspected community onset infection treated with intravenous antibiotics within 48 hours after admission.

In 1,299 patients with Sepsis-3 sepsis, the 28-day CFR was 12% (155/1,299), and among those with infection but not Sepsis-3 sepsis it was 2% (18/773). **Fig 1**.

The Sepsis-3 group of 1,299 patients consisted of subgroups with varying 28-day CFRs. One subgroup of 393 patients (30%) also had severe sepsis and a 28-day CFR of 26%. Another subgroup of 173 patients (13%) obtained a ≥2-point increase in the SOFA score by addition of 1-point organ dysfunction values only. The 28-day CFR was 2.9% (5/173). When adjusted for age, there was no statistical difference in the 28-day CFR between these patients and those having infection but not Sepsis-3 sepsis. Of these 173 patients, 27 also had severe sepsis. Among those 27 with severe sepsis were 4 of the 5 deaths. All 4 had elevated lactate levels >4 mmol/L. In the remaining 760 patients with Sepsis-3 sepsis, (1,299-393-173+27) the 28-day CFR was 7%. **Fig 2**, **Table 2**.

**Fig 2.**
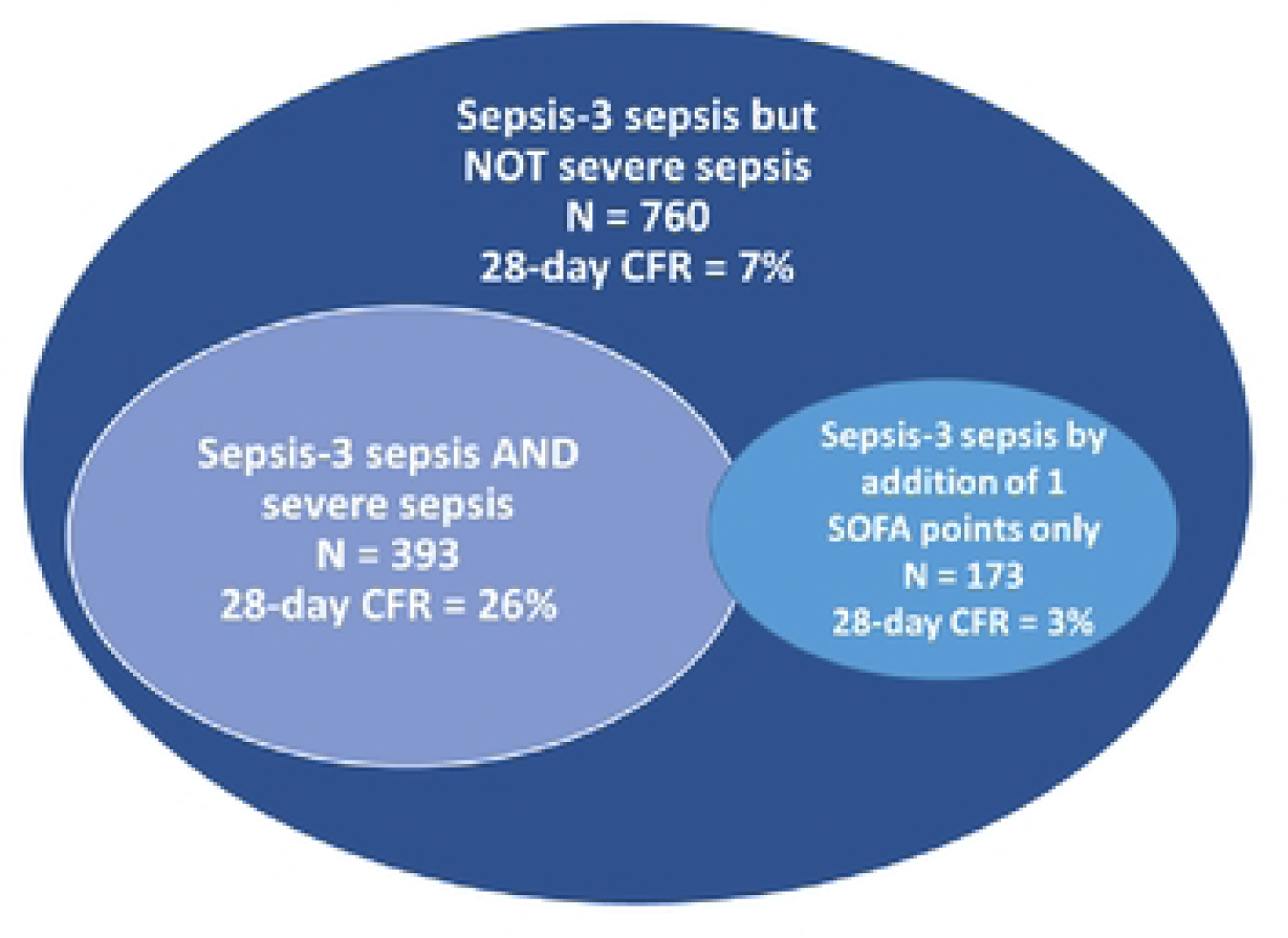
The distribution of subgroups with 28-day CFRs within a cohort of 1,299 patients with Sepsis-3 sepsis. Among the 173 who got a Sepsis-3 diagnosis by addition of 1-point changes in the SOFA score only, 27 also had severe sepsis.

**Table 2.**
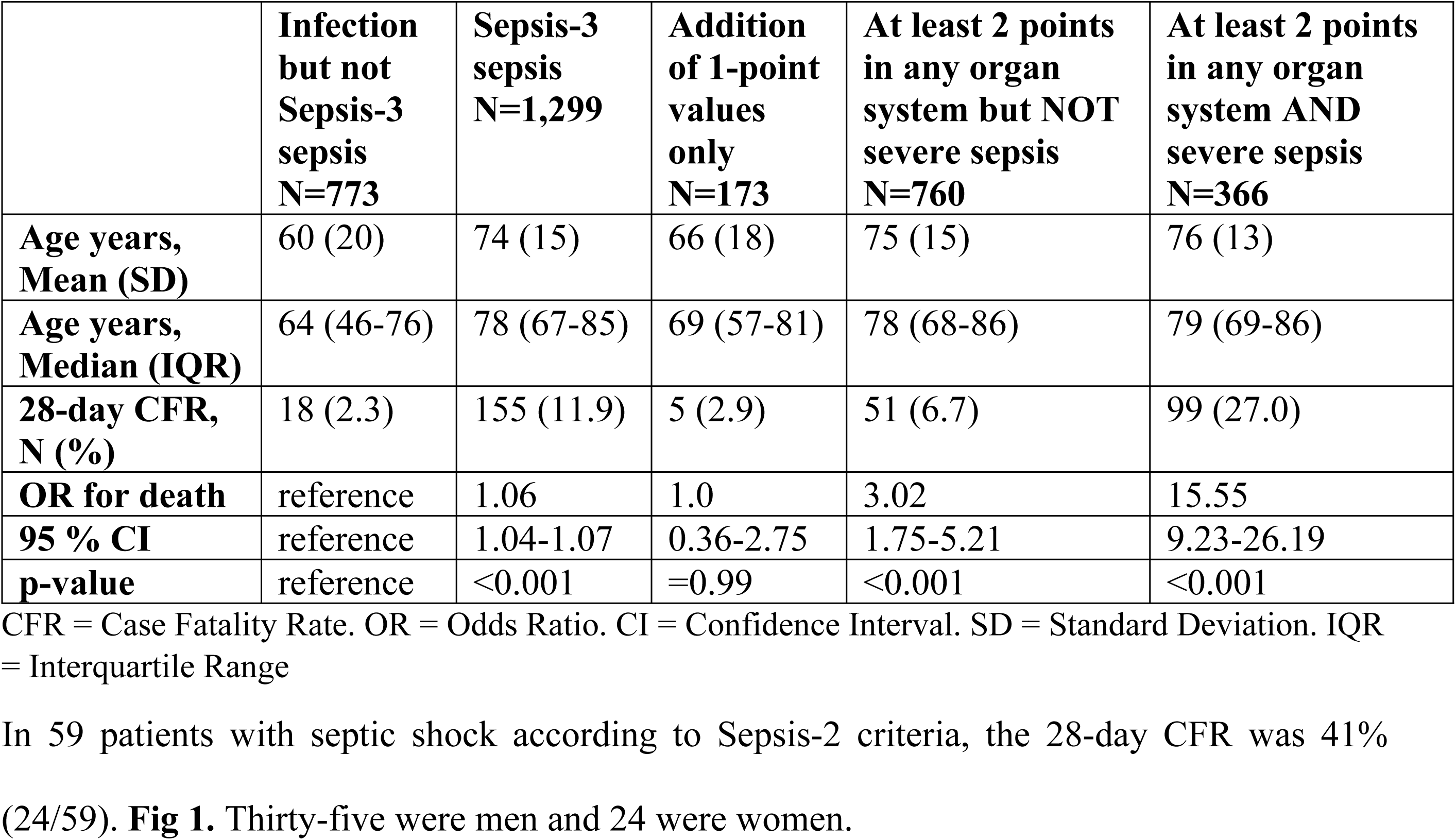
The odds ratio for 28-day CFR in subgroups of 1,299 patients with Sepsis-3 sepsis and for patients with infection but not Sepsis-3 sepsis. Adjusted for age by logistic regression analysis.

In 59 patients with septic shock according to Sepsis-2 criteria, the 28-day CFR was 41% (24/59). **Fig 1**. Thirty-five were men and 24 were women.

### Risk factors for 28-day case fatality in severe sepsis

Out of 429 patients with severe sepsis, 25% (107/429) died within 28 days. In the univariate analysis, age ≥85 years, respiratory dysfunction, renal dysfunction and low temperature on arrival were the strongest risk factors for case fatality. Notably, bacteremia did not turn out as a risk factor for case fatality. Since the p-value was >0,2 it is not in the table.

In the multivariate regression analysis, only age group >85 years, age group 75-84 years, renal dysfunction, cerebral dysfunction (RLS), elevated CRP level, and low temperature on arrival, remained as independent risk factors for case fatality. **Table 3**.

**Table 3.** Risk factors for 28-day case fatality in 429 patients with community onset severe sepsis.

| Variable | Univariate analysis<br>OR with 95% CI | p-value | Multivariate<br>analysis<sup>1</sup><br>OR with 95% CI | p-value |
| --- | --- | --- | --- | --- |
| **Age group, years** |  |  |  |  |
| <65 | reference |  |  |  |
| 65-74 | 2.31 (0.96-5.57) | 0.062 | 1.63 (0.41-6.46) | 0.490 |
| 75-84 | 3.47 (1.52-7.94) | 0.003 | 4.08* (1.10-15.15) | 0.036 |
| ≥85 | 5.21 (2.30-11.82) | <0.001 | 8.23* (2.18-31.0) | 0.002 |
| **Any co-morbidity** | 4.09 (1.23-13.61) | 0.022 | 1.27 (0.30-5.34) | 0.740 |
| **Respiratory dysfunction** | 2.57 (1.63-4.05) | <0.001 | 1.70 (0.76-3.81) | 0.199 |
| **Renal dysfunction** | 3.69 (2.21-6.14) | <0.001 | 3.20* (1.44-7.10) | 0.004 |
| **Hematologic dysfunction** | 1.96 (1.03-3.74) | 0.040 | 0.81 (0.28-2.33) | 0.698 |
| **Cerebral dysfunction** | 2.12 (1.34-3.36) | 0.001 | 2.04 (0.60-6.91) | 0.253 |
| **Hepatic dysfunction** | 5.50 (1.29-23.45) | 0.021 | 1.17 (0.12-11.27) | 0.890 |
| **RLS** | 2.21 (1.35-3.36) | 0.002 | 5.37* (1.45-19.90) | 0.012 |
| **Temperature** | 0.65 (0.53-0.80) | <0.001 | 0.66* (0.49-0.91) | 0.011 |
| **Saturation** | 0.97 (0.95-0.99) | 0.005 | 0.99 (0.95-1.02) | 0.486 |
| **History of fever/rigors** | 0.62 (0.40-0.97) | 0.036 | 1.08 (0.54-2.16) | 0.827 |
| **Hemoglobin** | 0.99 (0.98-0.998) | 0.020 | 0.99 (0.98-1.01) | 0.386 |
| **Lymphocytes** | 1.06 (0.97-1.16) | 0.190 | 1.09 (0.85-1.39) | 0.500 |
| **NLCR** | 0.99 (0.97-1.003) | 0.133 | 0.99 (0.97-1.01) | 0.163 |
| **CRP** | 1.003 (1.001-1.005) | 0.002 | 1.005* (1.002-1.008) | 0.001 |
| **INR** | 1.19 (0.93-1.15) | 0.157 | 1.01 (0.71-1.45) | 0.945 |
| **Lactate** | 1.11 (1.02-1.23) | 0.023 | 1.12 (0.95-1.32) | 0.162 |

<sup>1</sup>Multivariate logistic regression analysis with 28-day case fatality rate as outcome. RLS, Reaction Level Scale. NLCR, Neutrophil to lymphocyte count ratio. CRP, C-reactive protein. INR, International Normalized Ratio.

### Risk factors for 28-day case fatality in sepsis-3 sepsis

Out of 1,299 patients with sepsis-3 sepsis, 155 (12%) died within 28 days. In the multivariate regression analysis, only age group 75-84 years, >85 years, respiratory dysfunction, renal dysfunction, CRP, and low temperature on arrival, remained as independent risk factors for case fatality. Age ≥85 years, respiratory dysfunction, renal dysfunction, and CRP, were the variables with the highest statistical significance. **Table 4**.

**Table 4.**
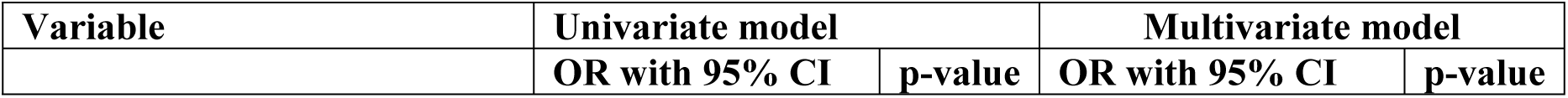
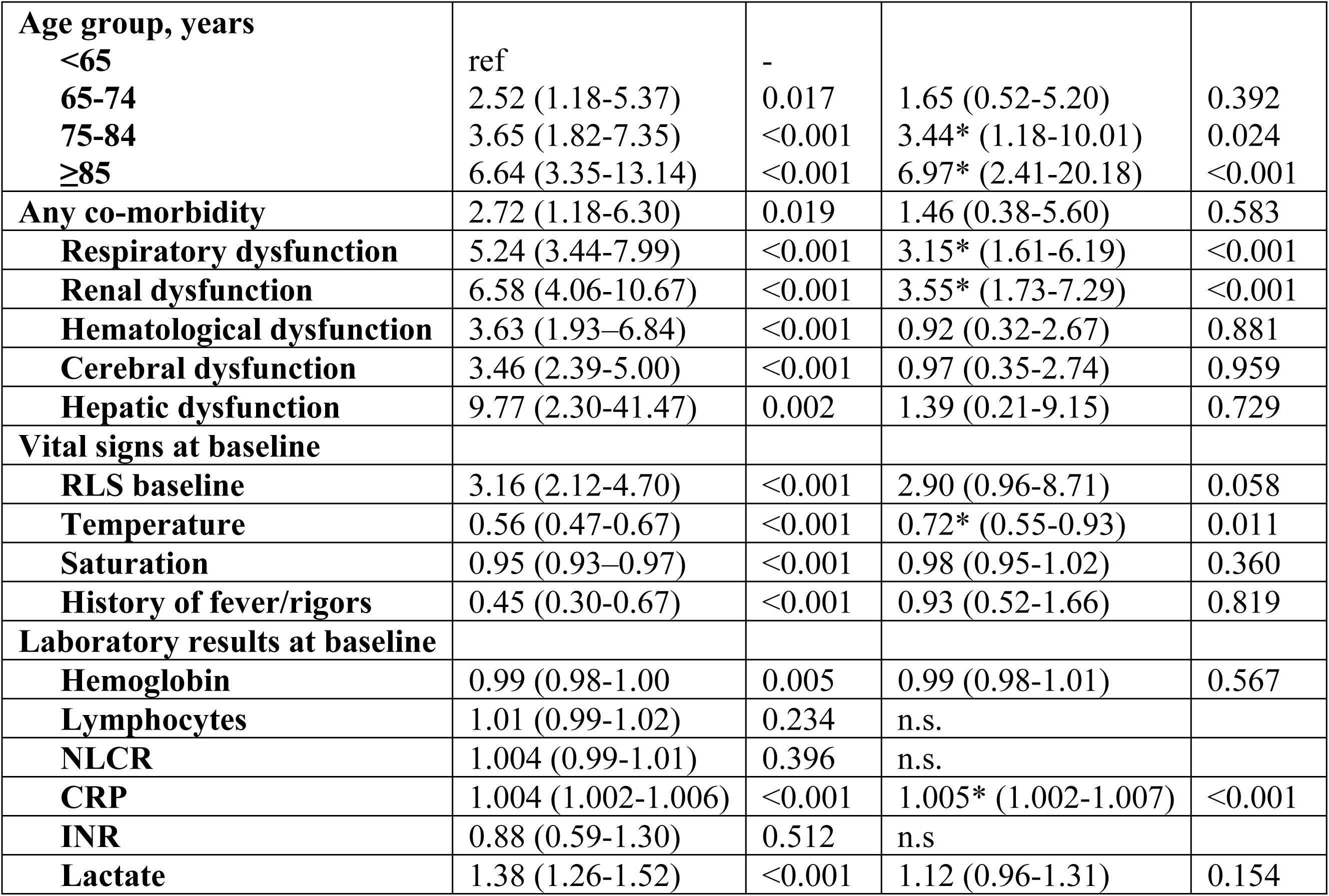
Risk factors for 28-days case fatality in 1,299 patients with sepsis-3 sepsis.

| Variable | Univariate model |  | Multivariate model |  |
| --- | --- | --- | --- | --- |
|  | OR with 95% CI | p-value | OR with 95% CI | p-value |
| **Age group, years** |  |  |  |  |
| **<65** | ref | - |  |  |
| **65-74** | 2.52 (1.18-5.37) | 0.017 | 1.65 (0.52-5.20) | 0.392 |
| **75-84** | 3.65 (1.82-7.35) | <0.001 | 3.44* (1.18-10.01) | 0.024 |
| **≥85** | 6.64 (3.35-13.14) | <0.001 | 6.97* (2.41-20.18) | <0.001 |
| **Any co-morbidity** | 2.72 (1.18-6.30) | 0.019 | 1.46 (0.38-5.60) | 0.583 |
| **Respiratory dysfunction** | 5.24 (3.44-7.99) | <0.001 | 3.15* (1.61-6.19) | <0.001 |
| **Renal dysfunction** | 6.58 (4.06-10.67) | <0.001 | 3.55* (1.73-7.29) | <0.001 |
| **Hematological dysfunction** | 3.63 (1.93-6.84) | <0.001 | 0.92 (0.32-2.67) | 0.881 |
| **Cerebral dysfunction** | 3.46 (2.39-5.00) | <0.001 | 0.97 (0.35-2.74) | 0.959 |
| **Hepatic dysfunction** | 9.77 (2.30-41.47) | 0.002 | 1.39 (0.21-9.15) | 0.729 |
| **Vital signs at baseline** |  |  |  |  |
| **RLS baseline** | 3.16 (2.12-4.70) | <0.001 | 2.90 (0.96-8.71) | 0.058 |
| **Temperature** | 0.56 (0.47-0.67) | <0.001 | 0.72* (0.55-0.93) | 0.011 |
| **Saturation** | 0.95 (0.93-0.97) | <0.001 | 0.98 (0.95-1.02) | 0.360 |
| **History of fever/rigors** | 0.45 (0.30-0.67) | <0.001 | 0.93 (0.52-1.66) | 0.819 |
| **Laboratory results at baseline** |  |  |  |  |
| **Hemoglobin** | 0.99 (0.98-1.00) | 0.005 | 0.99 (0.98-1.01) | 0.567 |
| **Lymphocytes** | 1.01 (0.99-1.02) | 0.234 | n.s. |  |
| **NLCR** | 1.004 (0.99-1.01) | 0.396 | n.s. |  |
| **CRP** | 1.004 (1.002-1.006) | <0.001 | 1.005* (1.002-1.007) | <0.001 |
| **INR** | 0.88 (0.59-1.30) | 0.512 | n.s. |  |
| **Lactate** | 1.38 (1.26-1.52) | <0.001 | 1.12 (0.96-1.31) | 0.154 |

### Age and sex

In patients with severe sepsis or septic shock, the 28-day CFR increased more than 10-fold from 3-36% between the youngest and the oldest age group. In patients with Sepsis-3 sepsis, the 28-day CFR also increased 10-fold, from 2-20%, with increasing age group. In the age group 18-49 years, the 28-day CFR was 2%. In the age-group 50-64 it was 5%, in the age-group 65-74 it was 9%, in the age-group 75-84 it was 12% and in those >85 years it was 20%. **Fig 3**. For number of patients in each group, see S3 Table. There was no statistically significant difference in 28-day CFR between men and women in any age group.

**Figure 3.**
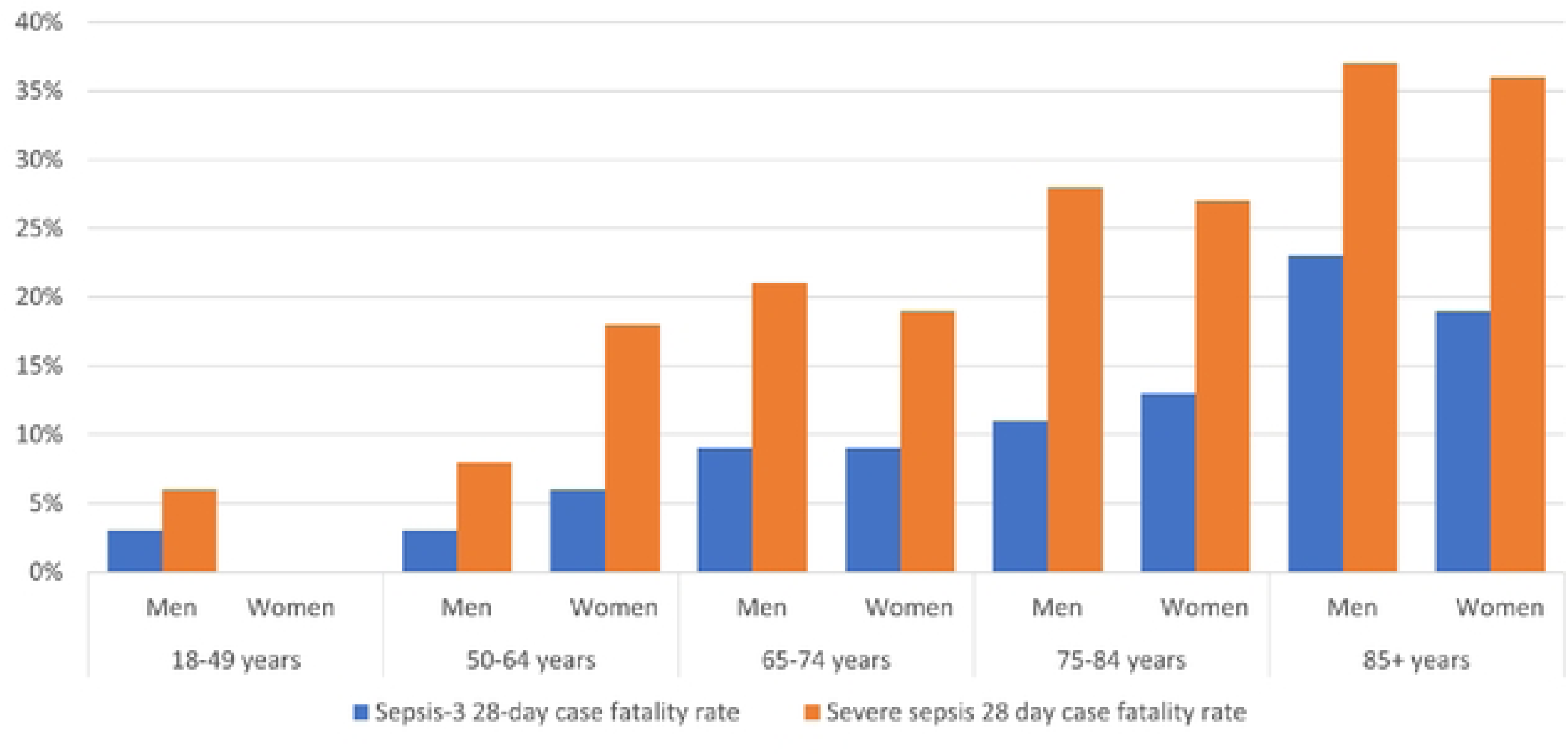
The 28-day case fatality rate related to age and sex in 1,299 patients having Sepsis-3 sepsis and 429 patients having severe sepsis. For numbers, see S3 Table.

### Appropriate antibiotic treatment

Considering the suspected focus of infection on admission, more than 98% of all patients were evaluated as having received appropriate empirical antibiotic treatment. In 952/2,072 (46%) patients with infection, a bacterial diagnosis was verified by culture. Of the 952 patients, 760 (80%) received appropriate etiological antibiotic treatment and 192 did not. The 28-day CFR was 7.8% and 5.7% respectively. The difference was not statistically significant (p-value 0,34), not even when adjusted for age (p-value 0.27).

### Time to appropriate antibiotic treatment

The 28-day case fatality rates in relation to time to start of antibiotic treatment is shown in **Table 5**. None of the differences in the 28-day CFRs are statistically significant. This data was not adjusted for age groups.

**Table 5.** The 28-day case fatality rate in relation to time to start of antibiotic treatment.

|  | Severe sepsis<br>N=429 | Infection but not<br>severe sepsis<br>N=1,643 | Sepsis-3 sepsis<br>N=1,299 | Infection but not<br>Sepsis-3 sepsis<br>N=773 |
| --- | --- | --- | --- | --- |
| Time to start of<br>antibiotics, hours | 28-day CFR, %<br>(N) | 28-day CFR, %<br>(N) | 28-day CFR, %<br>(N) | 28-day CFR, %<br>(N) |
| 0-0.99 | 23.1 (21) | 4.9 (8) | 14.5 (29) | 0 (0) |
| 1-1.99 | 20.6 (27) | 2.2 (10) | 8.9 (35) | 1 (2) |
| 2-3.99 | 33.0 (36) | 3.6 (22) | 12.8 (53) | 1.6 (5) |
| 4-11.99 | 21.8 (12) | 5.6 (16) | 10.5 (20) | 5.4 (8) |
| >12 | 25.6 (11) | 8.5 (10) | 17.5 (18) | 5.3 (3) |
| All | 24.9 (107) | 4.0 (66) | 11.9 (155) | 2.3 (18) |

There was a tendency towards shorter median length of hospital stay (LoS) for all patients with infection who received appropriate antibiotic treatment within 12 hours after admission. The difference was most pronounced in patients with severe sepsis, 11 versus 8 days. The calculations included only patients who survived 28 days but were not adjusted according to age group. **Table 6**.

**Table 6.**
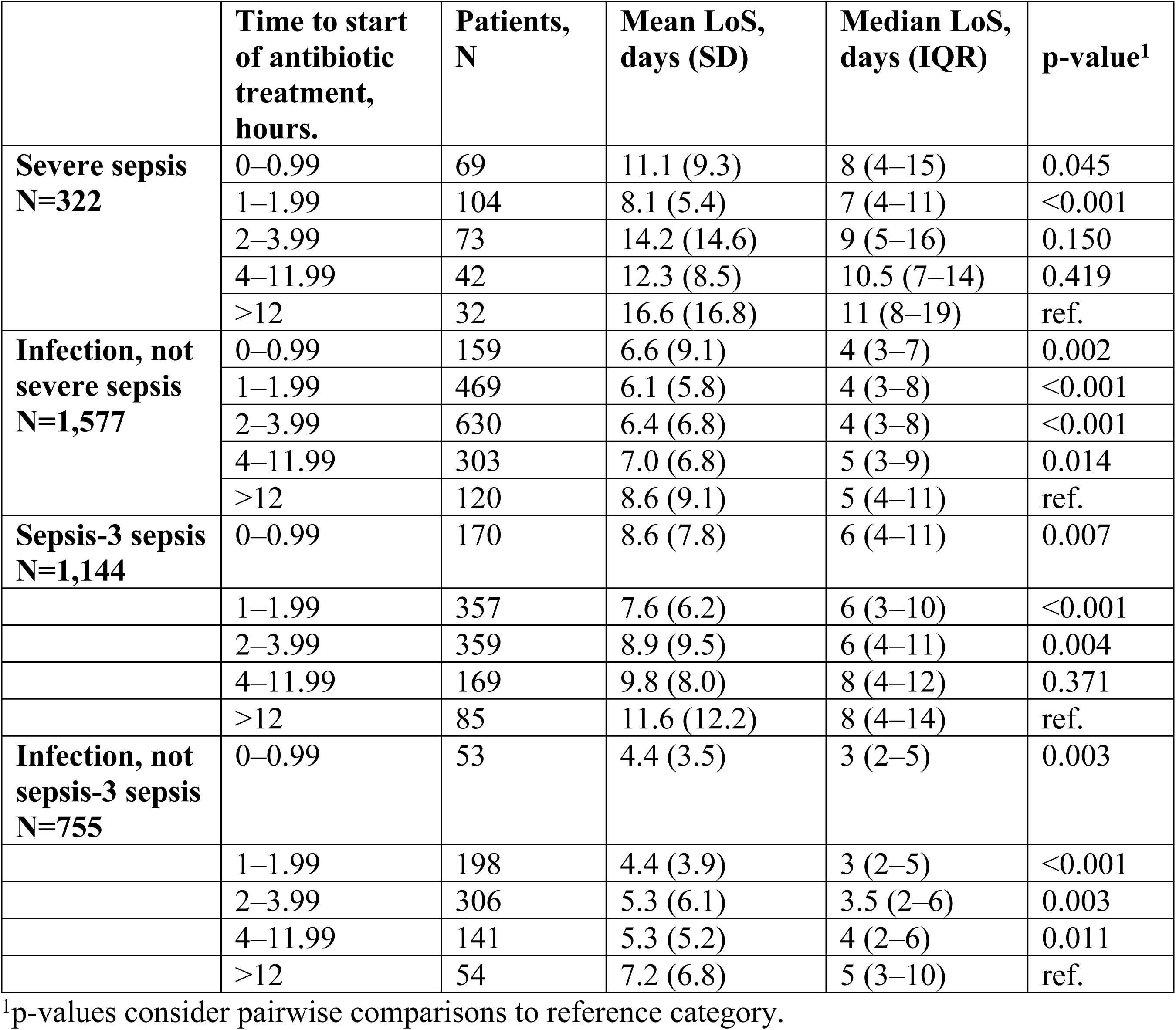
Length of hospital stay, LoS, (days) related to time to start of antibiotic treatment in patients with community onset infection.

|  | Time to start of antibiotic treatment, hours. | Patients, N | Mean LoS, days (SD) | Median LoS, days (IQR) | p-value<sup>1</sup> |
| --- | --- | --- | --- | --- | --- |
| **Severe sepsis**<br>**N=322** | 0–0.99 | 69 | 11.1 (9.3) | 8 (4–15) | 0.045 |
|  | 1–1.99 | 104 | 8.1 (5.4) | 7 (4–11) | <0.001 |
|  | 2–3.99 | 73 | 14.2 (14.6) | 9 (5–16) | 0.150 |
|  | 4–11.99 | 42 | 12.3 (8.5) | 10.5 (7–14) | 0.419 |
|  | >12 | 32 | 16.6 (16.8) | 11 (8–19) | ref. |
| **Infection, not**<br>**severe sepsis**<br>**N=1,577** | 0–0.99 | 159 | 6.6 (9.1) | 4 (3–7) | 0.002 |
|  | 1–1.99 | 469 | 6.1 (5.8) | 4 (3–8) | <0.001 |
|  | 2–3.99 | 630 | 6.4 (6.8) | 4 (3–8) | <0.001 |
|  | 4–11.99 | 303 | 7.0 (6.8) | 5 (3–9) | 0.014 |
|  | >12 | 120 | 8.6 (9.1) | 5 (4–11) | ref. |
| **Sepsis-3 sepsis**<br>**N=1,144** | 0–0.99 | 170 | 8.6 (7.8) | 6 (4–11) | 0.007 |
|  | 1–1.99 | 357 | 7.6 (6.2) | 6 (3–10) | <0.001 |
|  | 2–3.99 | 359 | 8.9 (9.5) | 6 (4–11) | 0.004 |
|  | 4–11.99 | 169 | 9.8 (8.0) | 8 (4–12) | 0.371 |
|  | >12 | 85 | 11.6 (12.2) | 8 (4–14) | ref. |
| **Infection, not**<br>**sepsis-3 sepsis**<br>**N=755** | 0–0.99 | 53 | 4.4 (3.5) | 3 (2–5) | 0.003 |
|  | 1–1.99 | 198 | 4.4 (3.9) | 3 (2–5) | <0.001 |
|  | 2–3.99 | 306 | 5.3 (6.1) | 3.5 (2–6) | 0.003 |
|  | 4–11.99 | 141 | 5.3 (5.2) | 4 (2–6) | 0.011 |
|  | >12 | 54 | 7.2 (6.8) | 5 (3–10) | ref. |

<sup>1</sup>p-values consider pairwise comparisons to reference category.

### Temperature

Low temperature on arrival was an independent risk factor for 28-day case fatality regardless of sepsis category or not, but most pronounced in those having severe sepsis. **Table 3 and 4.**

### Treatment in the intensive care unit

Out of 2,072 patients with infection of any severity, 170 (8%) were treated in the intensive care unit (ICU). The 28-day CFR was 21% (36/170). Severe sepsis or septic shock was diagnosed in 122/170. The 28-day CFR was 21% (25/122). Sepsis-3 sepsis was diagnosed in 163/170. The 28-day CFR was 22% (36/163).

### The accumulated case fatality rates up to 10 years after the first admission

The accumulated 1-year mortality rate for all 2,072 patients with infection was 23% (482/2,072), being up to twice as high for any age group as the 28-day CFR. The 1-year mortality in either the severe sepsis or the Sepsis-3 sepsis group was more than twice as high in each year group compared to those who had infection but not severe sepsis or Sepsis-3-sepsis. **Fig 4**. For patients with septic shock, the 1-year mortality was 64%.

**Figure 4.**
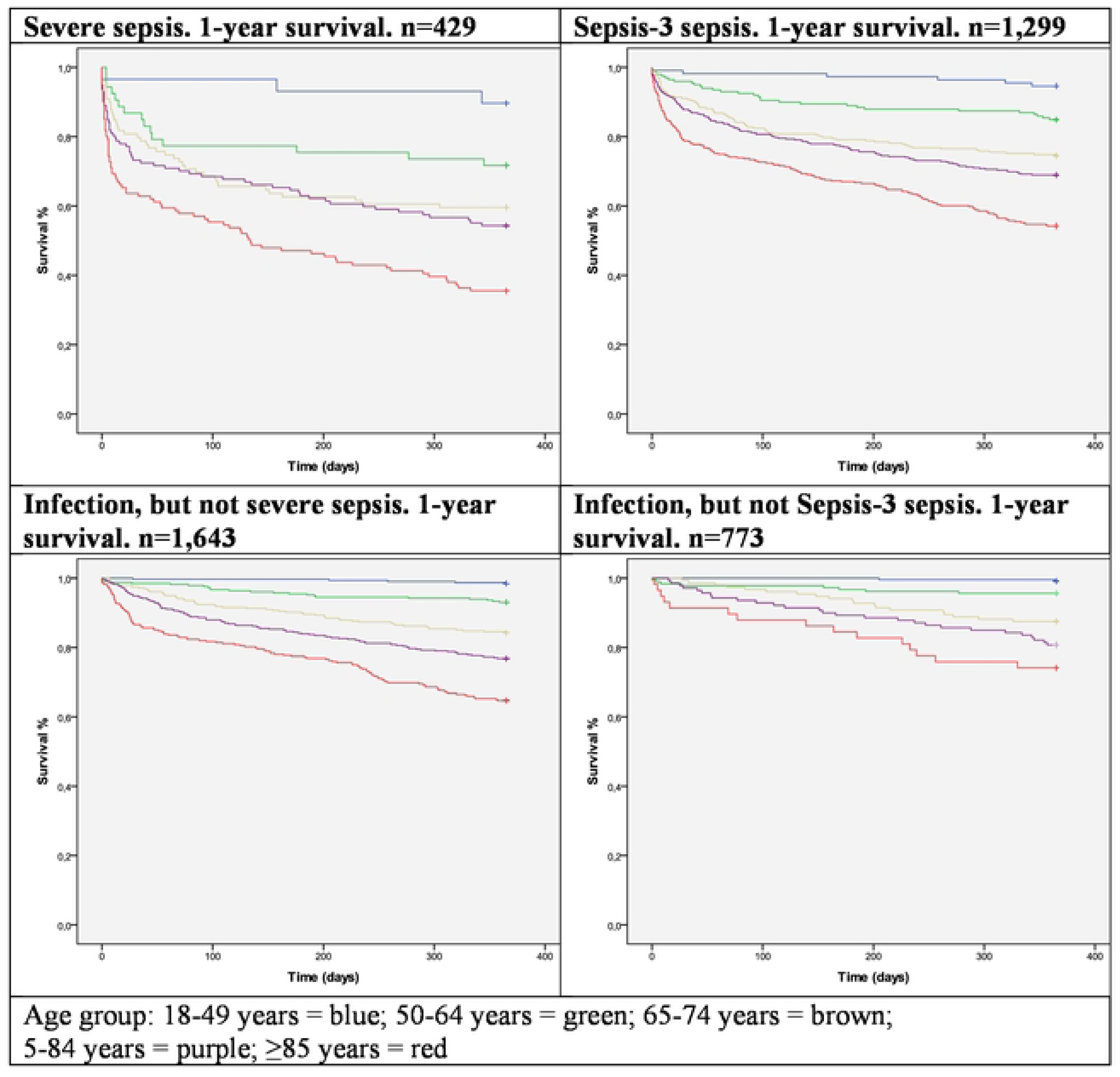
Accumulated 1-year mortality in different age-groups in patients with either severe sepsis or Sepsis-3 sepsis or not.

These differences remained during the whole follow up period of 10 years after first admission and were more pronounced in those with severe sepsis, regardless of age group. S1 Fig.

### Bacteremia

In 283 patients with a clinically relevant bacteremia the 28-day CFR was 12% (35/283). The 28-day CFR increased with increasing age in those having severe sepsis as well as in those having Sepsis-3 sepsis. **Table 7, 8.** Most of the 28-day case fatalities were seen in the 75-84 and ≥85-year age groups, 76% (22/29) among those with severe sepsis and 74% (23/31) among those with Sepsis-3 sepsis. The 28-day CFR was dependent on whether the patient had organ dysfunction or not. In patients with no organ dysfunction, it was not higher than in patients with no bacteremia. It also varied with etiology, but these figures are not adjusted for age. Bacteremia with *Staphylococcus aureus* had the highest 28-day CFR, 24%, followed by *non-pneumococcal streptococcus spp.,* 18%. For the most found bacteria, *Escherichia coli,* the 28-day CFR was 8%. The highest 28-day CFR was seen in those with *S. aureus* and severe sepsis, 53% (10/19).

**Table 7.** Patient characteristics and 28-day case fatality rates in 2,072 patients with infection depending on bacteremia or no bacteremia, severe sepsis or no severe sepsis. Due to small numbers, the 28-day CFRs according to etiology have not been adjusted for age.

|  | **Severe sepsis, N=429** |  | **No severe sepsis N=1,643** |  |
| --- | --- | --- | --- | --- |
|  | **Bacteremia N=123** | **No bacteremia N=306** | **Bacteremia N=160** | **No bacteremia N=1,483** |
| **Age, years, mean (SD)** | 77 (13) | 73 (16) | 72 (15) | 67 (19) |
| **Age, years, median (IQR)** | 80 (68-87) | 76 (67-85) | 76 (65-83) | 71 (57-81) |
| **28-day CFR, % (N)** | 24 (29/123) | 25 (78/306) | 4 (6/160) | 4 (60/1,483) |
| **28-day CFR per age group, % (N)** |  |  |  |  |
| **18-49 years, N=310** | 0 (0/5) | 4 (1/24) | 7 (1/14) | 0 (0/267) |
| **50-64 years, N=360** | 13 (2/16) | 14 (5/37) | 0 (0/26) | 1 (3/281) |
| **65-74 years, N=432** | 21 (5/24) | 20 (15/75) | 0 (0/36) | 2 (5/297) |
| **75-84 years, N=545** | 25 (9/36) | 29 (26/91) | 2 (1/53) | 5 (18/365) |
| **≥85 years, N=425** | 31 (13/42) | 39 (31/79) | 13 (4/31) | 13 (34/273) |
| **28-day CFR according to etiology, % (N)** |  |  |  |  |
| *E. coli* | 14 (6/44) | n.a. | 4 (2/54) | n.a. |
| *K. pneumoniae* | 13 (1/8) | n.a. | 0 (0/12) | n.a. |
| *S. aureus* | 53 (10/19) | n.a. | 6 (2/32) | n.a. |
| *S. pneumoniae* | 25 (3/12) | n.a. | 0 (0/8) | n.a. |
| *Non-pneumococcal streptococci* | 33 (6/18) | n.a. | 3 (1/29) | n.a. |

**Table 8.** Patient characteristics and 28-day case fatality rates in 2,072 patients with infection depending on bacteremia or no bacteremia, Sepsis-3-sepsis or not Sepsis-3 sepsis. Due to small numbers, the 28-day CFRs according to etiology have not been adjusted for age.

|  | **Sepsis-3 sepsis, N=1,299** |  | **Not sepsis-3 sepsis, N=773** |  |
| --- | --- | --- | --- | --- |
|  | **Bacteremia N=211** | **No bacteremia N=1,088** | **Bacteremia N=72** | **No bacteremia N=701** |
| **Age, years, mean (SD)** | 76 (14) | 74 (16) | 69 (16) | 60 (20) |
| **Age, years, median (IQR)** | 79 (68-86) | 77 (66-85) | 71 (62-81) | 63 (45-75) |
| **28-day CFR, % (n/N)** | 15% (31/211) | 11% (124/1088) | 6% (4/72) | 2% (14/701) |
| **28-day CFR per age group, % (n/N)** |  |  |  |  |
| **18-49 years, N=310** | 8% (1/12) | 1% (1/84) | 0% (0/7) | 0% (0/207) |
| **50-64 years, N=360** | 9% (2/23) | 4% (6/153) | 0% (0/19) | 2% (3/165) |
| **65-74 years, N=432** | 11% (5/44) | 8% (19/229) | 0% (0/16) | 1% (1/143) |
| **75-84 years, N=545** | 13% (9/69) | 12% (40/330) | 5% (1/20) | 5% (6/126) |
| ≥85 years, N=425 | 22% (14/63) | 20% (58/292) | 30% (3/10) | 7% (4/60) |
| 28-day CFR according to etiology, % (n/N) |  |  |  |  |
| *E. coli* (N=250) | 10% (7/73) | n.a. | 4% (1/23) | n.a. |
| *K. pneumoniae* (N=36) | 6% (1/18) | n.a. | 0% (0/2) | n.a. |
| *S. aureus* (N=105) | 29% (10/35) | n.a. | 14% (2/14) | n.a. |
| *S. pneumoniae* (N=73) | 19% (3/16) | n.a. | 0% (0/4) | n.a. |
| *Non-pneumococcal streptococci* (N=109) | 19% (6/32) | n.a. | 8% (1/13) | n.a. |

## Discussion

In this chart-based population study, the 28-day CFR in patients with severe sepsis was 25% and in patients with Sepsis-3 sepsis it was 12%. This is in line with the results in many previous studies. When comparing the groups, we found that the lower 28-day CFR in Sepsis-3 sepsis was achieved by adding to a group of patients with severe sepsis (393/429) and a high 28-day CFR (26%) a large group of equally old patients with a low 28-day CFR (6%). Thus, the lower 28-day CFR seen in Sepsis-3 sepsis was achieved by diluting the high 28-day CFR in patients having severe sepsis with a more than twice as large and equally old group of patients with a low 28-day CFR. Further, the 28-day CFR in the age-group 18-49 years was only 2%, not higher than in those with infection but not sepsis. In the age-groups 50-64 and 65-74 it was less than the 10% postulated by *Singer et.al.* for Sepsis-3 (5). It should be debated whether these patient groups with a 28-day CFR less than 10% really qualify for the Sepsis-3 definition of sepsis (“life-threatening organ dysfunction”) and if not the SOFA score should be revised, as suggested by *Szczeklik and Fronczek* (27). Especially the respiratory criterion for organ dysfunction should be stricter.

An increase of 1-point in the SOFA score can be obtained by mild organ dysfunction, in some cases for values that lie within the normal reference values for each parameter. In 173 patients with Sepsis-3 sepsis (13%), the diagnosis was achieved by addition of 1-point changes only. These patients had the same low 28-day CFR (3%) as those with infection but no organ dysfunction. This raises the question whether the 1-point values have any place in diagnosing organ dysfunction in Sepsis-3. From a clinical viewpoint it is difficult to appreciate that these patients should have “life-threatening organ dysfunction”.

The stricter criteria for respiratory dysfunction in the Swedish severe sepsis criteria from 2011 and in the Surviving Sepsis Campaign (SSC) criteria from 2012, seem to perform better in separating those having a truly “life-threatening organ dysfunction”, 25% 28-day CFR, from those having not, 4% 28-day CFR than the Sepsis-3 criteria. Though the 28-day CFR is high in patients with severe sepsis, it is so mainly because of the high CFR in the age groups >65 years old.

Increasing hospital mortality with increasing age in patients with severe sepsis was described in 2001 by Angus *et.al.* (17). Martin-Loechse *et.al.* found age to be an independent risk factor for case fatality, though only analyzing patients ≥65 years of age and only two age groups (19). Martin *et.al.* found age >65 years to be an independent risk factor for death from severe sepsis (18). In our population, both age groups 75-84 years and >85 years were independent risk factors for case fatality. We did not specifically study why there is an increase in CFRs with increasing age but believe that age is a surrogate marker for an ageing immune system, immunosenescence and reduced reserve capacity in organ functions. This view is to some extent supported by the fact that the average number of co-morbidities did not increase with increasing age-group (8). Interestingly, patients under the age of 50 years rarely died from community onset severe sepsis or Sepsis-3 sepsis. This is important knowledge but not so much highlighted in the literature. Though our cohort may be too small for such a conclusion, it is well in line with our more than 30-year experience from serving this population. In sepsis studies, case fatality rates should always be accounted for according to age or age groups.

Mortality rates depending on sex have varied in previous studies. We found a higher incidence of severe sepsis in men than in women (8) but no difference between the sexes in the 28-day CFR. Thus, men seem to be more prone to develop severe sepsis, but when severe sepsis is manifest, there is no difference in the 28-day CFR between the sexes. This supports the findings in the nationwide study on severe sepsis in the US by Angus *et.al.* in 2001 (20) but differs from the seasonal excess mortality seen for example in the Covid-19 pandemic where the case fatality rates among men are higher than in women, as described by *Nielsen et.al*. (28).

Bacteremia is a serious condition given the high CFRs in population studies, 10-13% (14, 29). In our study it was 12%. However, an increased CFR was seen only in those having organ dysfunction. Using severe sepsis criteria, only 43% of patients with bacteremia had organ dysfunction. Using Sepsis-3 criteria it was 75%. In non-sepsis patients the CFRs were not higher in patients with bacteremia than in those with infection but no bacteremia. Increasing age was also a significant risk factor for a high 28-day CFR. Another important factor affecting the CFR in bacteremia is the bacterial species. Thus, *E. coli* bacteremia is associated with a lower overall CFR (8%) (30) than *S. aureus* (20%) (31). Our study confirmed this. The highest CFR was seen in patients with *S. aureus* bacteremia and severe sepsis, 53%, compared to patients with *E. coli* bacteremia and severe sepsis, 14%. Most population studies on bacteremia are register-based, with no information about organ dysfunction. In a chart-based study like this, we see that it is not bacteremia per se that is associated with a high 28-day CFR, but whether the patient has organ dysfunction or not.

Is time to appropriate antibiotic treatment important for survival? In patients with septic shock, it probably is, as well as in certain cases of severe sepsis, but in this cohort, it could not be demonstrated. This may seem reasonable, since the median age of sepsis patients is high, and many have significant co-morbidities. In two studies from southern Sweden by Rosenqvist *et.al.* on a subgroup of patients with severe sepsis, not even appropriate antibiotic treatment within one hour after arrival affected the in-hospital mortality (32, 33). In a meta-analysis of studies on patients with bacterial infections, Naucler *et. al*. found that apart from patients with septic shock and acute bacterial meningitis, in-hospital mortality was not increased if antibiotic treatment was started up to 12 hours after hospital admission (34). In one study, Rhee et. al. estimated that only 11 out of 300 sepsis-associated deaths may have been prevented by more timely antibiotic treatment (35). This raises the question whether mortality is the most relevant outcome measure in sepsis studies. Most sepsis patients survive, and to them morbidity may be a better outcome measure, though difficult to define. In this study, we could see a tendency towards shorter length of hospital stay (LoS) in all patient groups with infection if appropriate empirical antibiotic treatment was started within two to four hours after arrival. This effect was most pronounced in patients with severe sepsis.

Temperature on arrival has been shown to correlate inversely to in-hospital mortality in a cohort of Swedish sepsis patients treated in the ICU, Sundén-Cullberg *et.al.* (22). In this population we found this to be true for all patients with infection, though most pronounced in those with severe sepsis.

The long-term CFR was higher in those having severe sepsis, versus those having not, and higher in those having Sepsis-3 sepsis versus those having not. Thus, this population-based study verifies the results of previous studies done on different cohorts reaching the same conclusion (36).

## Conclusion

This is to our knowledge the only chart-based prospective and consecutive study on the outcome of community onset severe sepsis in adults in a whole population.

In this single center study on community onset sepsis

- Sepsis-2 criteria with stricter criteria for respiratory dysfunction are better for separating those with a truly life-threatening organ dysfunction from those having not, than the Sepsis-3 criteria.
- there is a ten-fold difference in the 28-day CFR between the youngest and the oldest age groups.
- age is an independent risk factor for case fatality in sepsis.
- the lower CFR seen in Sepsis-3 sepsis is due to a dilution of a group of patients with severe sepsis and a high CFR with a twice as large, equally old, group of patients with a low CFR.
- patients who obtain ≥2 points in the SOFA score by addition of 1-point values only, have the same low CFR as patients who do not have Sepsis-3 sepsis.
- the 28-day CFR in patients with bacteremia, or “blood stream infection”, is related to organ dysfunction, not to bacteremia *per se*.
- time to appropriate antibiotic treatment does not affect the 28-day CFR but tends to shorten the LoS for all patient groups with infection, mainly for those with severe sepsis.
- temperature on arrival is inversely associated with the 28-day CFR for all groups with infection.
- the long-term mortality in patients with sepsis is increased for each age group up to ten years after the first episode and is more pronounced in those with severe sepsis.

## Data Availability

All original data will be made available if the article is accepted. They were published after the first article was accepted in your journal in 2019, but currently I don't know where they can be found.

